# Set-based rare variant association tests for biobank scale sequencing data sets

**DOI:** 10.1101/2021.07.12.21260400

**Authors:** Wei Zhou, Wenjian Bi, Zhangchen Zhao, Kushal K. Dey, Karthik A. Jagadeesh, Konrad J. Karczewski, Mark J. Daly, Benjamin M. Neale, Seunggeun Lee

## Abstract

UK Biobank has released the large-scale whole-exome sequencing (WES) data, but the best practices remain unclear for rare variant tests, and an existing approach, SAIGE-GENE, can have inflated type I error rates with high computation costs. Here, we propose SAIGE-GENE+ with greatly improved type I error control and computational efficiency compared to SAIGE-GENE. In the analysis of UKBB WES data of 30 quantitative and 141 binary traits, SAIGE-GENE+ identified 551 gene-phenotype associations. In addition, we showed that incorporating multiple MAF cutoffs and functional annotations can help identify novel gene-phenotype associations.

## Main

UK Biobank (UKBB) recently released the whole-exome sequencing (WES) data of most of their participants^1^, allowing for studying rare variant associations for complex traits and diseases. However, the best practices remain unclear for set-based rare variant tests in large-scale biobanks. A common practice is to test for associations with all rare (minor allele frequency, MAF ≤1%,) loss-of-function (LoF) and missense variants, but this approach can lose power if association signals are enriched in very rare variants or certain functional annotation classes. To improve power, researchers can restrict the test for rarer variants such as variants with MAF ≤0.1% or MAF ≤0.01%. Another approach is incorporating functional annotations. Since multiple variant sets with different MAF cutoffs and functional annotations exist, tests should be done multiple times for each gene or region and results should be aggregated using the minimum p-value or the Cauchy combination^2,3^.

Currently, SAIGE-GENE^4^ is the only method developed to conduct the non-burden type set-based rare variant tests, such as SKAT^5^ and SKAT-O^6^, for binary phenotypes with unbalanced case-control ratios in biobank-scale data. For example, in our evaluation, the most recent set-based test, STAAR^2^, cannot control for type I error rates in the presence of case-control imbalance (**Supplementary Figure 1**). Burden tests can be done with single variant test methods and implemented in software packages including REGENIE2^7^, but the Burden test can suffer from a lower power compared to the SKAT-O test that is more robust to proportion of causal variants and direction of causal effects^6^, which we also observed in simulation studies **(Methods, Supplementary Note A, Supplementary Figure 2)**. SAIGE-GENE has been recently used for exome-wide association analysis for 3,700 phenotypes of 281,850 individuals in the UKBB WES data^8^. In the evaluation of the method using the UKBB WES data with 160K white British samples, we have found that all the tests (Burden, SKAT and SKAT-O) in SAIGE-GENE performed well when testing all rare variants with MAF 1% (**Figure 1A**), but inflation was observed in SKAT and SKAT-O tests in SAIGE-GENE when restricting to variants with MAF ≤0.1% or MAF≤0.01% and the case-control rates were more unbalanced than 1:30 (**Figure 1A, Supplementary Figure 3)**. To examine whether the inflation is due to inflated type I error rates or polygenicity of the phenotypes, we carried out type I error simulation studies with various case-control ratios (**Supplementary Note B** and **Methods**), and observed the same inflation (**Supplementary Figure 4**). This suggests that SKAT and SKAT-O in SAIGE-GENE can suffer from inflated type I error rates.

**Figure 1.**
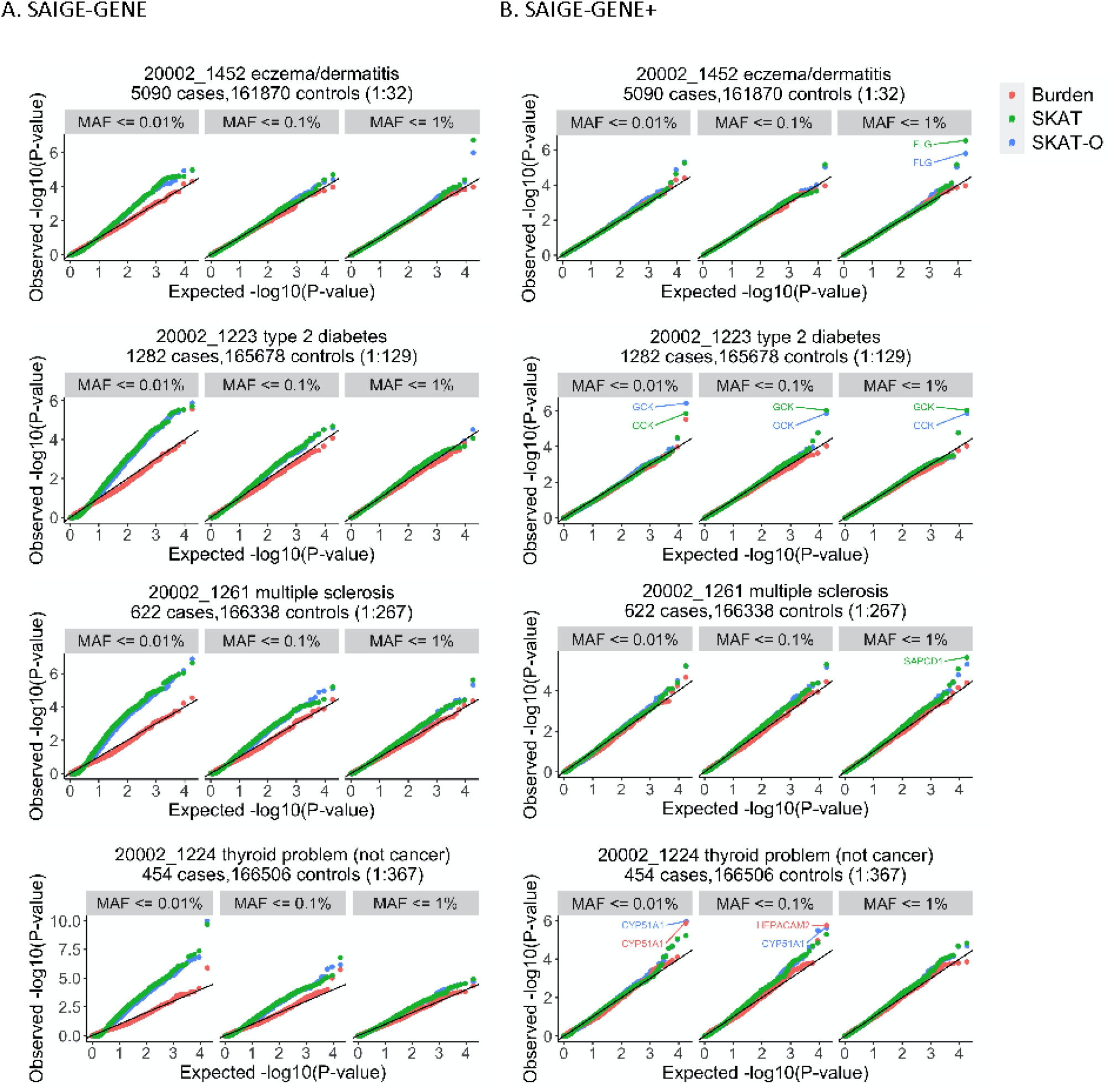
Quantile-quantile (Q-Q) plots for Burden, SKAT^3^ and SKAT-O^4^ for four exemplary binary phenotypes in the UKBB WES data using A. SAIGE-GENE and B. SAIGE-GENE+. The tests were performed for 18,372 genes with missense and loss-of-function (LoF) variants with three different maximum MAF cutoffs: 1%, 0.1%, and 0.01%. Names of genes reaching the exome-wide significant threshold (p-value < 2.5×10^−6^) in SAIGE-GENE+ are annotated in the plots.

In addition, the computation cost is not low enough to test for the multiple variant sets. For example, to test the largest gene (*TTN*) with 16,227 variants in the UKBB WES data with 3 different MAF cutoffs (MAF≤1%, ≤0.1%, ≤0.01%) and 3 different annotations (LoF only, LoF+missense, and LoF+missense+synonymous), SAIGE-GENE required 164 CPU hours and 65 Gigabytes (Gb) of memory (**Supplementary Table 1**).

To address these issues, we propose SAIGE-GENE+. Although SAIGE-GENE uses various approaches, including saddlepoint approximation and exact resampling, to adjust for unbalanced case-control ratios, these approaches cannot fully address the imbalance and sparsity in the data (**Figure 1A, Supplementary Figure 4A**). In order to reduce the sparsity, prior to the set-based association tests, SAIGE-GENE+ collapses ultra-rare variants with MAC ≤10 in the “absence and presence” way that has been commonly adapted in the association analysis of ultra-rare variants^9,10^ by assuming those variants have the same effect direction on the testing phenotype (**Methods)**. We observed that the inflation of SKAT and SKAT-O has been substantially reduced in SAIGE-GENE+ and all tests have well controlled type I errors in both simulated (**Supplementary Figure 4B**) and the UKBB WES data (**Figure 1B**) for four exemplary phenotypes with case-control ratios 1:32 to 1:267. The genomic control inflation factors also became closer to one (**Supplementary Figure 3)**.

Collapsing ultra-rare variants in SAIGE-GENE+ decreases the number of variants in each gene or region (**Supplementary Figure 5**), leading to reduced computation time and memory usage (**Figure 2A, Supplementary Table 1, Supplementary Figure 6**). To further reduce computational cost, SAIGE-GENE+ reads in genotypes or dosages for all genetic markers only once, followed by conducting multiple association tests corresponding to different MAF cutoffs and annotations. The computation time of SAIGE-GENE+ for conducting all Burden, SKAT, and SKAT-O was 1,407 times decreased (9,851 mins vs. 7 mins) and the memory usage dropped from 65Gb to 2.1Gb compared to SAIGE-GENE when testing the largest gene *TTN (*16,227 LoF+missense+synonymous variants) for its association with the basal metabolic rate (**Supplementary Table 1**). To test 18,372 genes with 150,000 samples using SKAT-O tests with three MAF cutoff 1%, 0.1%, and 0.01% and three different variant annotations: LoF only, LoF+missense, and LoF+missense+synonymous, SAIGE-GENE+ costs 78.6 CPU hours (18.8 CPU hours for fitting the null mixed model using a full GRM as Step 1 and 59.8 CPU hours for association tests as Step 2) and maximum 4.8Gb memory (4.8Gb for Step 1 and 2 Gb for Step 2) (**Supplementary Tables 2 and 3, Supplementary Figure 7**). In addition, when a sparse GRM instead of a full GRM is used in Step 1 in SAIGE-GENE+, the time cost and memory usage dramatically dropped for fitting the null mixed model (< 1 min and 0.61Gb) (**Supplementary Table 2, Supplementary Note C**). We also compared the computation time of SAIGE-GENE+ and REGENIE2 (**Supplementary Table 2** and **3, Supplementary Note D, Supplementary Figure 7**).

**Figure 2.**
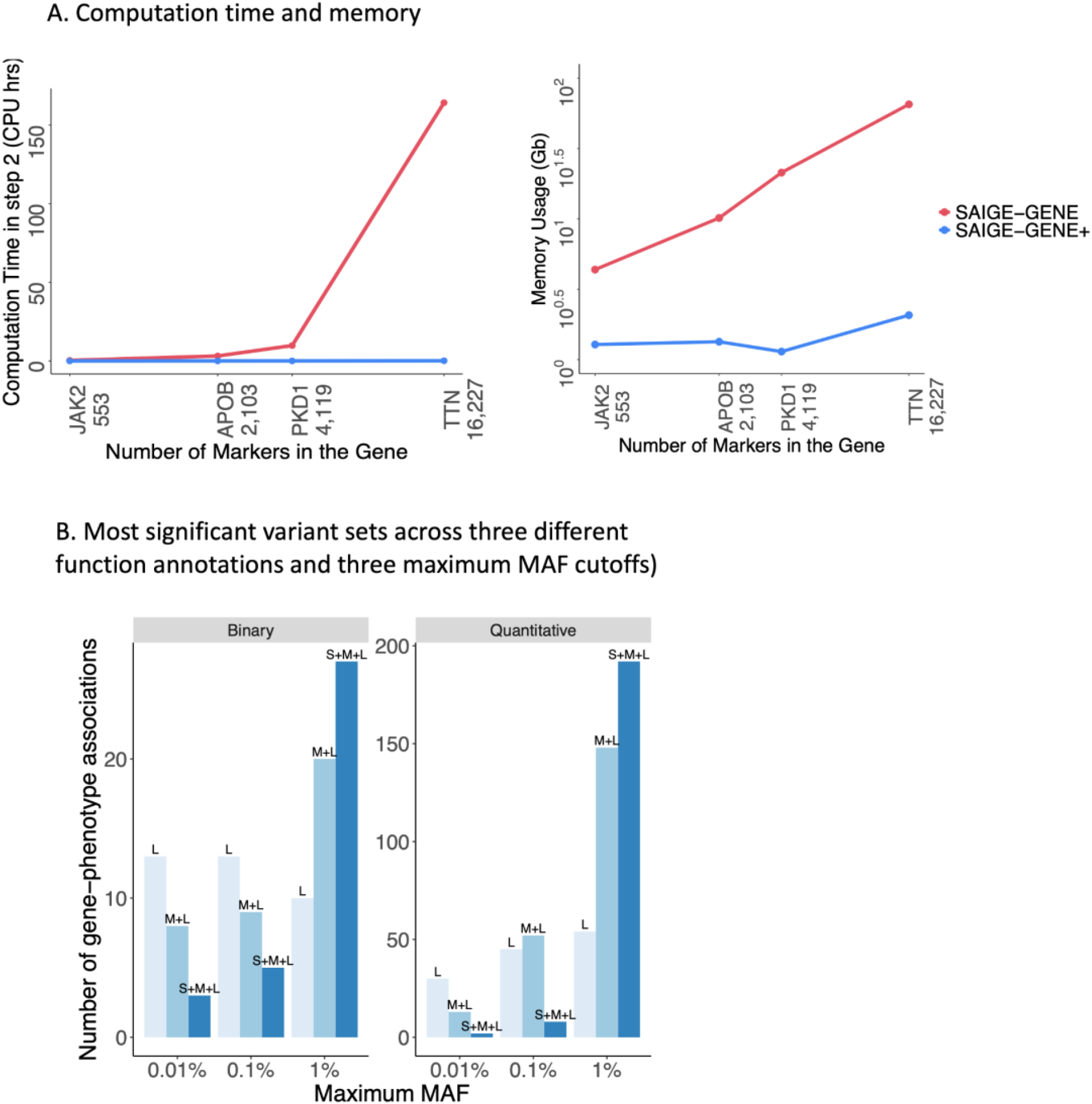
Performance of SAIGE-GENE+ in UK Biobank WES data. **A. Computation time and memory of the gene-based tests (Step2, See Method) in SAIGE-GENE and SAIGE-GENE+ for four example genes with different numbers of variants**. The SKAT-O tests were conducted with three maximum MAF cutoffs: 1%, 0.1%, and 0.01% and three variant annotations: LoF only, LoF + missense, and LoF + missense +synonymous and combined using the Cauchy combination. The plots are in the log10-log10 scale. The details of the numbers and genes are presented in the **Supplementary Table 1**. **B. Most significant variant sets across the three different MAF cutoffs: 1%**, *0*.**1%, and 0.01% and three functional annotations: LoF (L) only, LoF + missense (M+L), and LoF + missense +synonymous (S+M+L)**. Distribution of variant sets with the smallest p-values, among 551 significant gene-phenotype associations identified by SAIGE-GENE+ in the analyses of 30 quantitative traits and 141 binary traits in the UKBB WES data.

By collapsing ultra-rare variants, SAIGE-GENE+ can have more significant p-values for several well-known gene-phenotype associations than SAIGE-GENE, even though the latter has inflated type I errors. We applied SAIGE-GENE and SAIGE-GENE+ to 37 self-reported binary phenotypes in the UKBB WES data using three different maximum MAF cutoffs: 1%, 0.1%, and 0.01% to 18,372 gene including all missense and LoF variants. We have observed 27 SKAT-O tests with more significant p-values in SAIGE-GENE+ than in SAIGE-GENE (**Supplementary Table 4**). For example, *BRCA2* for breast cancer with MAF ≤0.1% had a p-value 7.62×10^−8^ in SAIGE-GENE+ and 1.65×10^−3^ in SAIGE-GENE. Similarly, we observed the gene *GCK* for diabetes with maximum MAF ≤0.1% had a more significant p-value (1.22×10^−13^) in SAIGE-GENE+ than in SAIGE-GENE (p-value = 4.06×10^−6^). More detailed discussion can be found in **Supplementary Note E**.

We evaluated the powers of SAIGE-GENE+ and SAIGE-GENE through extensive simulation studies based on the real genotypes of 10 genes in the UKBB WES data (**Supplementary Table 5, Supplementary Note A, Methods**) for simulated binary traits. Three scenarios with different settings of proportions of causal variants across the multiple functional annotations and different settings of absolute effect sizes for causal variants were used. For each scenario, two settings of effect directions were used: 1. All causal variants increased disease risk; 2. 100% of LoF, 80% of missense, and 50% of synonymous causal variants increased disease risk and the other causal variants decreased disease risk (**Supplementary Table 6**). The prevalence of the binary phenotypes was set to be 10%, under which both SAIGE-GENE and SAIGE-GENE+ have well controlled type I error rates for Burden, SKAT, and SKAT-O tests. We observed that in all scenarios, SAIGE-GENE+ had higher or similar empirical power than SAIGE-GENE (**Supplementary Figure 8)** with the increased median Chi-square statistics (**Supplementary Table 7**). Our results also demonstrated as previously reported that the SKAT-O test can have higher power than the Burden test with more significant p-values and higher median Chi-square statistics (**Supplementary Figure 2, Supplementary Table 7**), while the Burden test p-values from SAIGE-GENE+ are highly concordant to p-values from REGENIE2 (Pearson’s correlation R^2^ = 0.99 for -log_10_(p-value)) (**Supplementary Figure 9**). In addition, the simulation results suggested incorporating multiple functional annotations (LoF, LoF+Missense, LoF+Missense+Synonymous) and maximum MAF cutoffs (0.01%, 0.1%, and 1%) can have an increased power compared to only using a single maximum MAF cutoffs (1%) on one set of function annotation (LoF+Missense+Synonymous) (**Supplementary Figure 10, Supplementary Table 7)**.

We applied SAIGE-GENE+ to analyze 18,372 genes in the UKBB 160K white British sample data using three different MAF cutoffs: 1%, 0.1%, and 0.01% and three different variant annotations: LoF only, LoF+missense, and LoF+missense +synonymous for 30 quantitative and 141 binary traits. The nine SKAT-O test p-values were then combined using Cauchy combination or the minimum p-value approach (**Methods)**. 465 gene-phenotype associations were significant at the exome-wide significance threshold (p-value ≤2.5×10^−6^) for 27 quantitative traits (**Supplementary Table 8**) and 86 gene-phenotype associations for 51 binary traits (**Supplementary Table 9**). Since the expected number of p-values < 2.5×10^−6^ under no association across all the phenotypes is 7.85, the false discovery rate is 0.014. Known genes *BRCA1, BRCA2, CHEK2, PALB2*, and *SAMHD1* were significant for breast cancer, *CNTNAP3B, CDKN2A* and *MITF* for melanoma, *IL33* for asthma, *GCK* for type 2 diabetes, and *LDLR* for high cholesterol and ischemic heart disease. For quantitative traits, *MC4R* and *GIPR* were significant for body mass index, *CETP, LIPG*, and *LPL* for the HDL cholesterol, *LDLR* and *PCSK9* for LDL, and *MEPE* for heel bone mineral density. We also identified potentially novel gene-phenotype associations. For binary traits, a pancreatic cancer susceptibility gene *NOC2L*^*11*^ was significant for hypovolemia (p-value = 9.68×10^−7^), *CYP21A2*, known for the 21-hydroxylase deficient congenital adrenal hyperplasia (CAH)^12^, for allergy/adverse effect of penicillin (p-value = 3.63×10^−7^). Also for quantitative traits, *CHEK2*, known to be associated with breast cancer and sex hormone-binding globulin measurement^13^, was significant for age at menopause (p-value = 4.51×10^−17^), *IGLL5*, which encodes an immunoglobulin lambda-like polypeptide, for the lymphocyte count (p-value = 1.50×10^−12^), and *NANOG*, which mediates germline development^14^ and is highly expressed in embryonic carcinoma^15^, for age at first live birth (p-value = 2.01 ×10^−6^).

Using lower MAF cutoffs may help identify novel associations in which the association signals are highly enriched in the rarer variant. For example, the association between *PDCD1LG2*, which encodes Programmed Cell Death 1 Ligand, and chronic lymphoid leukemia became significant when tests were restricted to variants with MAF ≤0.01% and 0.1% (p-value= 7.5×10^−7^) compared to testing all variants with MAF ≤1% (p-value = 5.4×10^−4^) (**Supplementary Table 10)**. The underlying reason for this observation could be that the associations are largely enriched in the rarer variants in the gene, e.g. the most significant variant is the low frequency variant rs7854303 (9:5557672, MAF=3.4×10^−4^) (see the Phe Web browser). Using a higher MAC cutoff 1% will include many non-causal variants, which decreases the power of the set-based association tests. In addition, including lower MAF cutoff sets helped to replicate known associations including *MLH1* for colorectal cancer and *CDKN2A* for melanoma (**Supplementary Table 10)**. Due to the multiple comparison burden, including lower MAF cutoff sets can make marginally significant associations non-significant. For 141 binary phenotypes, 17 out of 92 (18.4%) associations were further identified with lower MAF cutoff sets, while 9 (9.8%) associations became non-significant (**Supplementary Figure 11A, Supplementary Table 10**). For 30 quantitative traits, 28 out of 465 (6%) associations were further identified, while 53 (11.4%) associations became non-significant (**Supplementary Figure 11A, Supplementary Table 11**), suggesting that restricting association tests to rarer variants has a gain for binary phenotypes. In functional annotation categories, 184 associations were identified when the tests were conducted on LoF variants only, including LoF+missense sets identified 299 additional associations, and when LoF+missense+synonymous sets were also included, 91 more associations were identified (**Supplementary Figure 11B**). These results are consistent with the observation in the simulation studies that empirical power increased when incorporating multiple functional annotation combinations and maximum MAF cutoffs (**Supplementary Figure 10**). We also investigated among the 551 significant gene-phenotype associations, which variant set had the smallest p-value (**Figure 2B**). Interestingly, among sets with MAF ≤ 0.01%, LoF variant sets generally had the smallest p-values, while when the MAF cutoff ≤ 1%, LoF+missense+synonymous sets generally had the smallest p-values.

In summary, our results demonstrated that by incorporating multiple MAF cutoffs and functional annotations the exome-wide rare-variant association tests can help identify novel gene-phenotype associations and our SAIGE-GENE+ can facilitate this. Moreover, SAIGE-GENE+ has an option to allow users to fit the null generalized linear mixed model with a sparse genetic relationship matrix (GRM) to further reduce the computation burden (**Supplementary Note C and Supplementary Figure 12 to 14**).

## Methods

### Collapsing ultra-rare variants

Ultra-rare variants with MAC ≤10 were collapsed to a single marker (**Supplementary Figure 15**). More specifically, if no per-marker weights are provided by the user, all ultra-rare variants will be collapsed to a new variant in the “absence and presence” way^9^. The dosage for each sample was assigned as the maximum dosage value among all ultra-rare variants carried by the sample, if any. Then the weights of the collapsed variant and non ultra-rare variants (MAC > 10) are calculated based on their MAF from beta distribution *Beta*(*MAF, a*_*1*_, *a*_*2*_). By default, same as the setting of the SKAT-O test, *a*_*1*_ = 1 and *a*_*1*_ = 25 are used. If the per-marker weights are provided by the user, the ultra-rare variants will be collapsed to a new variant whose dosages are the maximum values among the weighted dosages.

### Aggregating multiple tests

For each gene or region, p-values of multiple testing sets corresponding to multiple maximum MAF cutoffs and functional annotations were aggregated using the Cauchy combination^2,3^. Note that the Cauchy combination does not work when any p-value is 1. Thus, we used the minimum p-value with the Bonferroni correction to combine multiple tests when at least one test had p-value = 1.

### Type I error evaluation

To evaluate the type I error control of SAIGE-GENE and SAIGE-GENE+, we simulated binary phenotypes under the null hypothesis of no genetic effects based on the observed genotypes by WES of the 166,955 samples with white British ancestry in the UK Biobank (**Supplementary Note B**). We conducted gene-based tests for 7,932 genes on the even chromosomes with missense and LoF variants using three different maximum MAF cutoffs: 1%, 0.1%, and 0.01%. In total, 158,640 gene-based tests were conducted for each maximum MAF cutoff for SAIGE-GENE and SAIGE-GENE+, respectively, and the Q-Q plots were shown in **Supplementary Figure 4**. Our simulation results suggest that SAIGE-GENE+ has well controlled type I errors with case-control ratios < 1:100 when testing variants with maximum minor allele frequency (MAF) = 0.01% (**Supplementary Figure 4B**).

We also evaluated the type I error control of SAIGE-GENE, SAIGE-GENE+ and STAAR (**Figure 1, Supplementary Figure 1**) in real data. We applied the methods to four exemplary self-reported binary phenotypes with various case control ratios in the UKBB WES data to 18,372 genes including all LoF and missense variants using three different maximum MAF cutoffs: 1%, 0.1%, and 0.01%. For STAAR, we used the relative coefficient cutoff 0.05 for the sparse GRM to fit the null models.

### Power evaluation

To evaluate the power of SAIGE-GENE+ and SAIGE-GENE, we simulated binary phenotypes based on the genotypes of ten genes in the WES data of 166,955 samples with white British ancestry in the UK Biobank (**Supplementary Table 5**). The selected genes showed significant gene-phenotype associations (**Supplementary Table 4**) and had wide ranges of the number of rare variants from 2901 (*APOB*) to 107 (*GPSM3*). The prevalence of the phenotype was set to be 10%, under which, both SAIGE-GENE and SAIGE-GENE+ have well controlled type I error rates for Burden, SKAT, and SKAT-O tests (**Supplementary Figure 3**). Three scenarios with different settings of proportions of causal variants and magnitudes of effect sizes for causal variants were used:1) low proportion of causal variants and low effect sizes, 2) low proportion of causal variants and high effect sizes, and 3) high proportion of causal variants and high effect sizes (**Supplementary Table 6**). More details about the simulation settings are described in **Supplementary Note A**. Our simulation results suggest that SAIGE-GENE+ has higher or similar empirical power than SAIGE-GENE (**Supplementary Figure 8** and **Supplementary Table 7**).

### UK Biobank WES data analysis

We applied SAIGE-GENE+ to analyze 18,372 genes in the UKBB WES data of 166,955 white British samples for 30 quantitative traits and 141 binary traits. Three different maximum MAF cutoffs: 1%, 0.1%, and 0.01% and three different variant annotations: LoF only, LoF + missense, and LoF + missense +synonymous were applied followed by aggregating the multiple SKAT-O tests using the Cauchy combination^2,3^ for each gene. Variants were annotated using ANNOVAR^16^. The LoF variants include those annotated as frameshift deletion, frameshift insertion, non-frameshift deletion, non-frameshift insertion, splicing, stop gain, and stop loss. Sex, age when attended assessment center, and first four PCs that were estimated using all samples with White British ancestry were adjusted in all tests. 250,656 pruned markers with MAF ≥ 1%, which were pruned from the directly genotyped markers using the following parameters, were used to construct GRM: window size of 500 base pairs (bp), step-size of 50 bp, and pairwise r2 < 0.2. We used the relative coefficient cutoff 0.05 for the sparse GRM for the variance ratio estimation after fitting the null models. The model was fitted with leave-one-chromosome-out (LOCO) to avoid proximal contamination.

### Computation cost evaluation

Benchmarking was performed on randomly sub-sampled UK Biobank WES data (up to 150,000 samples) with White British participants for glaucoma (1,741 cases and 162,408 controls). We report the medians of five runs for run times and memory with samples randomly selected from the full sample set using five different sampling seeds. SAIGE-GENE and SAIGE-GENE+ use a two-step approach. Step1 estimates the model parameters (i.e. variance component and fixed effect coefficients) in the null model and Step2 conducts set-based association tests. SAIGE-GENE+ runs Step 1 with all covariates as offset, which leads to a decrease of the computation time (**Supplementary Table 12**). We then compared computation time and memory usages of Step2 (**Figure 2A, Supplementary Table 1, Supplementary Figure 6**). Note that model parameters need to be estimated only once for each phenotype and can be used genome-wide regardless of MAF cutoffs and functional annotations. The computation cost of Step1 in SAIGE-GENE+ was given in **Supplementary Figure 7A** and **Supplementary Table 2**. SAIGE-GENE+ has an option to use a sparse genetic relationship matrix (GRM), which further reduces computation cost in Step1 (**Supplementary Note C**).

## Code and data availability

SAIGE-GENE+ is implemented as an open-source R package available at https://github.com/saigegit/SAIGE. SAIGE-GENE is available at https://github.com/weizhouUMICH/SAIGE/master. REGENIE (version 2.2.4) was downloaded from https://github.com/rgcgithub/regenie. STAAR (version 0.9.5) was downloaded from https://github.com/xihaoli/STAAR The PheWeb^17^ like visual server for 30 quantitative and 141 binary phenotypes of UK Biobank WES data analysis results are currently available at https://ukb-200kexome.leelabsg.org

## Data Availability

SAIGE-GENE+ is implemented as an open-source R package available at https://github.com/saigegit/SAIGE. SAIGE-GENE is available at https://github.com/weizhouUMICH/SAIGE/master. The summary statistics and QQ plots for 30 quantitative phenotypes and 141 binary phenotypes in UK Biobank by SAIGE-GENE+ are currently available for public download at
https://storage.googleapis.com/leelabsg/saige-gene/reformat_all_withPhenoDetails.txt

https://storage.googleapis.com/leelabsg/saige-gene/reformat_all_withPhenoDetails.txt

## Acknowledgments

This research has been conducted using the UK Biobank Resource under application number 45227. SL was supported by Brain Pool Plus (BP+, Brain Pool+) Program through the National Research Foundation of Korea (NRF) funded by the Ministry of Science and ICT (2020H1D3A2A03100666, S.L). WB and ZZ were supported by NIH R01 HG008773. WZ was supported by the National Human Genome Research Institute of the National Institutes of Health under award number T32HG010464. We thank Dr. Alkes Price for the constructive comments and suggestions.

## Author contributions

W.Z., W.B., Z.Z., and S.L. designed experiments. W.Z., W.B., Z.Z., performed experiments and analyzed the UK Biobank data. W.Z. implemented the software with input from W.B. and Z.Z.. Helpful advice was provided by K.K.D, K.A.J., K.J.K., B.M.N., and M.J.D.. W.Z., W.B., Z.Z., and S.L. wrote the manuscript with input from all co-authors.

