## Supplementary Note and Figures for "Set-based rare variant association tests for biobank scale sequencing data sets"

### A. Simulation studies for power evaluation

Phenotypes were simulated based on the real genotypes $G$ of randomly selected $L$= 30,000 LD-pruned ($r^2$ < 0.2) markers from the odd chromosomes with MAF $\geq$ 1% from the following logistic mixed model logistic mixed model $logit(\pi_{i0}) = \alpha_0 + X_{i1} + X_{i2} + \sum_{j=1}^{L} \hat{G}_{ij}\beta + \sum g_{iqk}\beta_{ci}$, where $\pi_{i0}$ is the probability for the $ith$ individual being a case given covariates and random effects, $\hat{G}_{ij}$ is the standardized genotype value for the $jth$ marker of $ith$ individual, and $\beta$ is the genetic effect size following $N(0, \tau/L)$, where $\tau = 1$, which is the variance component parameter. Two covariates, $X_{i1}$and $X_{i2}$, were simulated from Bernoulli(0.5) and N(0,1). The intercept $\alpha_0$ was determined by the prevalence 10%. In addition to the 30,000 variants to simulate random effects, we select 10 genes as causal genes to simulate phenotypes and evaluate power. $g_{iqk}$ is the genotype value for the $qth$ variant in the $kth$ gene of $ith$ individual, $\beta_{Ci}$ is the causal genetic effect sizes. Since functionally severe variants are likely to be causal variants with high effect sizes, we considered a different proportion of causal variants and effect sizes by the functional annotations.Two different settings of proportions of causal variants across the multiple functional annotations were used for variants that are not ultra-rare (MAC > 10): 1. 20% of LoF, 10% of missense, and 2% of synonymous and 2. 30% of LoF, 10% of missense, and 2% of synonymous. As functionally deleterious variants are more likely to be rarer, we assumed that the proportions of causal variants in the ultra-rare variants were 3 times higher than the numbers above (**Supplementary Table 6**). Two settings of the absolute effect sizes for causal missense and synonymous variants were used, $|0.25\log_{10}(\text{MAF})|$ and $|0.15\log_{10}(\text{MAF})|$, respectively, while the absolute effect sizes for causal LoF variants were set to be twice greater: $|0.5\log_{10}(\text{MAF})|$ and $|0.3\log_{10}(\text{MAF})|$. Two different settings of effect directions were used among causal variants, 1. all causal variants had the same effect direction 2. 100% of LoF, 80% of missense, and 50% of the synonymous variants increased disease risk while the other causal variants decreased disease risks. In the second setting (different association direction), since LoF variants are not likely to have different effect directions, we still assumed that the effect directions of LoF causal variants were the same. We repeated the simulations 100 times for different simulation scenarios, resulting in 1000 p-values (10 genes with each having 100 p-values) for each scenario.

### B. Simulation studies for type I error evaluation

The phenotypes were simulated based on real genotypes of randomly selected $L$= 30,000 LD-pruned ($r^2$ < 0.2) markers from the odd chromosomes with MAF $\geq$ 1% from the following logistic mixed model $logit(\pi_{i0}) = \alpha_0 + X_{i1} + X_{i2} + \sum_{j=1}^{L} \hat{G}_{ij}\beta$, where $\pi_{i0}$ is the probability for the $ith$ individual being a case given covariates $X_{i1}$and $X_{i2}$, and random effects, $\hat{G}_{ij}$. We followed the same scenarios to simulate covariates, random effects, and effect size $\beta$ as in power simulations. The intercept $\alpha_0$ was determined by given prevalence (i.e. case-control ratios). We repeated the simulation for 20 times for different disease prevalence: 0.3%, 1%, and 10%, respectively. For each phenotype set, a null logistic mixed model was fitted in Step 1 with covariates including the first 4 genetic principal components, which were estimated for all White-British participants in the UK Biobank, $X_1$ and $X_2$.

### C. Additional evaluation for set-based tests using the sparse GRM in the UK Biobank

A recently developed generalized linear mixed model method, fastGWA-GLMM[1], proposed using the sparse GRM to fit the null generalized linear mixed model for single-variants association tests in the UKBB.

In SAIGE-GENE+, we implemented this option to allow users to fit the null generalized mixed model using a sparse GRM. We applied this option to the UKBB WES data and compared the exome-wide association results to those using the full GRM. Three different coefficients of relationship cutoffs 0.05, 0.0884 (up to 3rd degree of relationship) and 0.176 (up to 2nd degree of relationship) have been applied to the sparse GRM (**Supplementary Figure 12**). We observed that the association p-values using the sparse GRM with different coefficients of relationship cutoffs are highly correlated with those using the full GRM across different binary traits (**Supplementary Figure 12**) and quantitative phenotypes (**Supplementary Figure 14**). Step 1 to fit the null model required a much smaller computation time when using the sparse GRM (**Supplementary Figure 7**). For example, with 150,000 samples, Step 1 required < 1 CPU min with sparse GRM (up to 3rd degree of relationship), while it took 11 CPU hours with full GRM estimated on-the-fly with 93511 genetic markers (**Supplementary Table 2**). Since the sample relatedness in the UKBB is modest, the computational performance gain of using the sparse GRM can be modest in data sets with widespread sample relatedness.

### D. Comparison with REGENIE

We observed that the Burden test p-values by SAIGE-GENE+ are highly concordant with the p-values by REGENIE2 (Pearson's correlation $R^2$ = 0.99 for $-\log_{10}$(p-value)) (**Supplementary Figure 9**). We also compared the empirical computation cost of SAIGE-GENE+ and REGENIE2[2]. In Step 1 for fitting null models, SAIGE-GENE+ with a full GRM was more efficient than REGENIE2 (**Supplementary Figure 7A and Supplementary Table 2**). Out of the five runs with 150,000 samples that were randomly sub-sampled from the UK Biobank WES data with White British participants for glaucoma (1,741 cases and 162,408 controls) from the UKBB, the median computation time for Step 1 is 11 CPU hours using SAIGE-GENE+ and 36.5 CPU hours using REGENIE2 and the median memory usage is 5.4 Gb in SAIGE-GENE+ and 7.3 Gb in REGENIE2**.** Moreover, when a sparse GRM instead of a full GRM is used in Step 1 in SAIGE-GENE+, the time cost and memory usage dramatically dropped (< 1 min and 0.61Gb)**.** In Step 2, similar computation cost was observed for the two methods for Burden tests **(Supplementary Figure 7B** and **Supplementary Table 3):** 8.8 CPU hours and 0.93Gb by REGENIE2 and 9.1 CPU hours and 0.97Gb by SAIGE-GENE+, which additionally output the p-values by the Cauchy combination. SAIGE-GENE+ conducts the SKAT-O test, while REGENIE2 conducts Burden tests only and does not allow for incorporating marker level weights. Although the SKAT-O tests in SAIGE-GENE+ required nearly 6-7x more computation time (60 CPU hours) and 2x more memory (2.0 Gb) (**Supplementary Figure 7B** and **Supplementary Table 3**), through simulation studies, we observed that SKAT-O tests have higher power than Burden tests in all different scenario (**Supplementary Table 6**) with more significant p-values (**Supplementary Figure 2**) and higher median Chi-square statistics (**Supplementary Table 7**).

#### E. SAIGE-GENE+ and SAIGE comparison in BRCA2 and GCK

*BRCA2* for breast cancer with MAF $\leq$ *0*.1% had p-value $7.62 \times 10^{-8}$ in SAIGE-GENE+ and $1.65 \times 10^{-3}$ in SAIGE-GENE. Similarly, we observed the gene *GCK* for diabetes with maximum MAF 0.1% had a more significant p-value ($1.22 \times 10^{-13}$) in SAIGE-GENE+ than in SAIGE-GENE (p-value = $4.06 \times 10^{-6}$).

In BRCA2-Breast Cancer, the associations are highly enriched in the ultra-rare LoF variants that tend to have the same effect directions, as is observed that the collapsed variant from 142 ultra-rare LoF variants had p-value $4.9 \times 10^{-22}$ ( https://ukb-200kexome.leelabsg.org/assoc/BRCA2/20001_1002). It is known that the Burden test is more powerful than the SKAT[3] test when most of the genetic variants in the test set are causal (having non-zero effects) with the same effect direction, whereas the SKAT test is more powerful when a small proportion of genetic variants are causal with inconsistent effect directions. Without collapsing in SAIGE-GENE, the Burden test p-value (0.000738) is more significant than the SKAT test p-value (0.114). But because only a small proportion of variants are causal, Burden test can still suffer from the low association power. After collapsing in SAIGE-GENE+, the SKAT test (p-value=$2.67\times10^{-8}$) had a more significant p-value than the Burden test (p-value = 0.00452). This could be because the association signal is largely contributed by the collapsed ultra-rare variants and the SKAT test is more robust to the large proportion of non-causal variants in the test sets. Similarly, it is observed that the association between the gene GCK and diabetes is driven by the collapsed ultra-rare variants https://ukb-200kexome.leelabsg.org/assoc/GCK/2443.

### F. Implementation of SAIGE-GENE+ to improve the computational efficiency

In order to further improve the computational efficiency, we utilized several approaches in the implementation of SAIGE-GENE+. 1. In Step 1 for fitting the null model, covariates are treated as offset which decreases the computation time (**Supplementary Table 12**). 2. When incorporating multiple MAF cutoffs and functional annotations, genotypes or dosages for all markers for each testing gene or region are read in at once and subsets are extracted for different tests. This largely reduces the redundancy for reading in genotype or dosages. 3. We implemented the Score tests and SPA tests in Rcpp, which reduces the overhead of switching between C++ and R. 4. For the tested set with $q$ markers, as illustrated in the SAIGE-GENE paper, the $q$ x $q$ matrix $R^{1/2}G'P_sGR^{1/2}$ is used to approximate the G'PG, whose eigenvalues are used to obtain the statistics of the SKAT test. To reduce memory usage, we divide the q markers to several blocks and store the submatrices that are required to approximate G'PG in the hard disk, and then compute the corresponding submatrix of G'PG using each pair of the blocks. This approach to save memory usage has been previously used in other programs, such as KING[4] for estimating sample relatedness.

**Supplementary Figure 1. Quantile-quantile plots for STAAR-O[5] tests p-values for four exemplary binary phenotypes with different case-control ratios in the UKBB WES data.** The tests were performed for 18,372 genes with missense and loss-of-function (LoF) variants with three different maximum MAF cutoffs: 1%, 0.1%, and 0.01%.

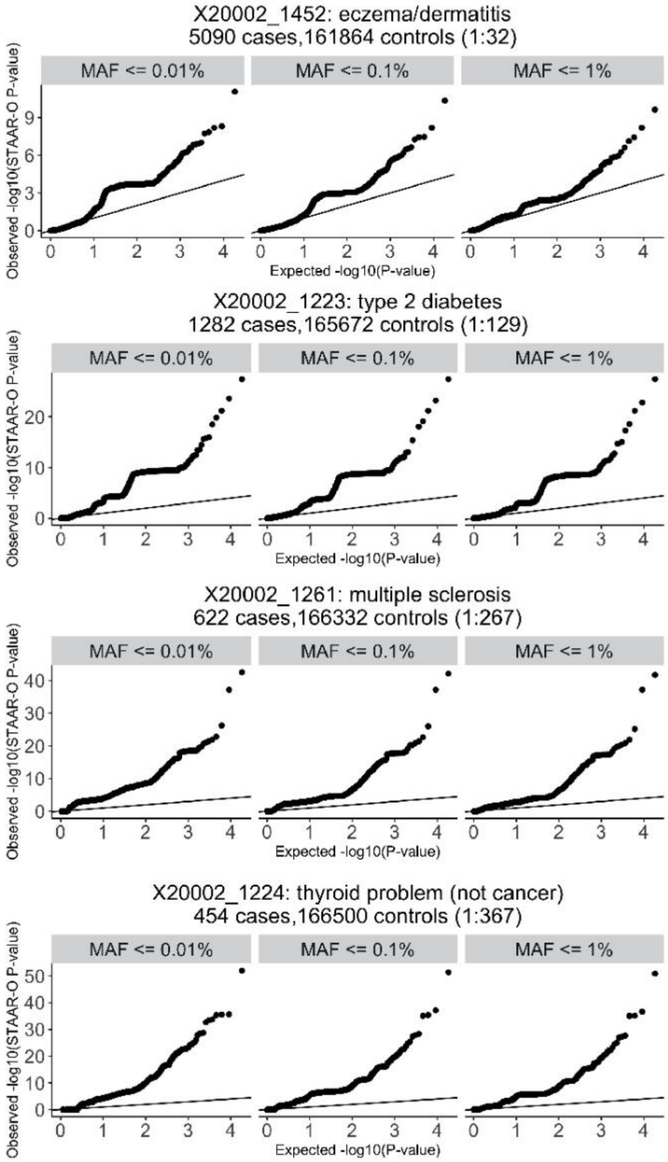

**Supplementary Figure 2. Scatter plots for association p-values of SKAT-O and Burden tests in the simulation studies.** Each plot is based on test results for 1,000 test sets (100 data sets, each of which includes 10 genes, see **Supplementary Table 5**). X-axis represents -log10 Burden test p-values, and Y-axis represents -log10 SKAT-O p-values. The line in each plot represents the 45-degree line, so the dots above the line indicate more significant p-values from SKAT-O than the Burden test. The details of different simulation settings are presented in **Supplementary Table 6.**

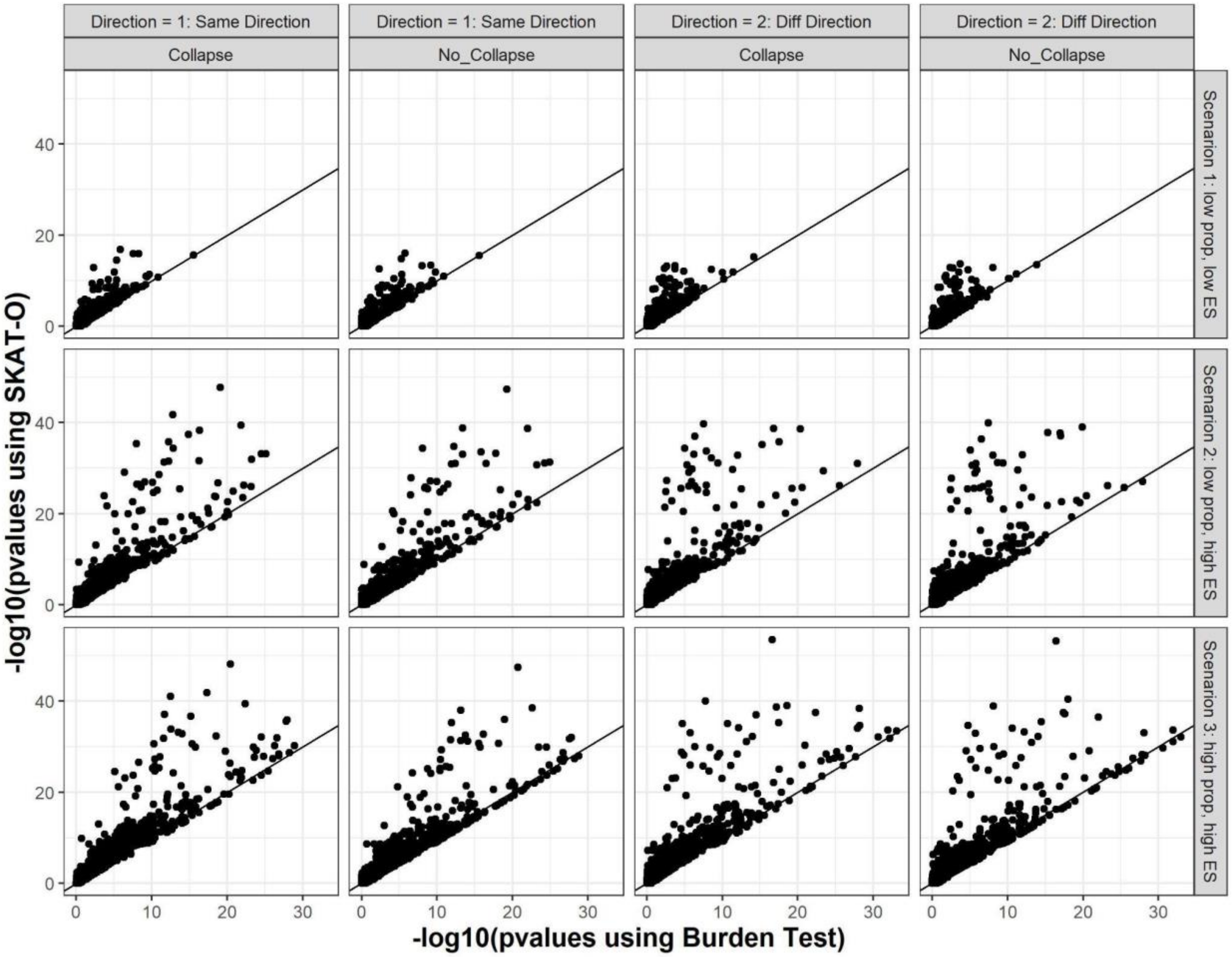

**Supplementary Figure 3.** The genomic control inflation lambda based on the 1st percentile against the disease prevalence for 24 binary phenotypes in UKBB for SAIGE-GENE and SAIGE-GENE+ using three different maximum MAF cutoffs.

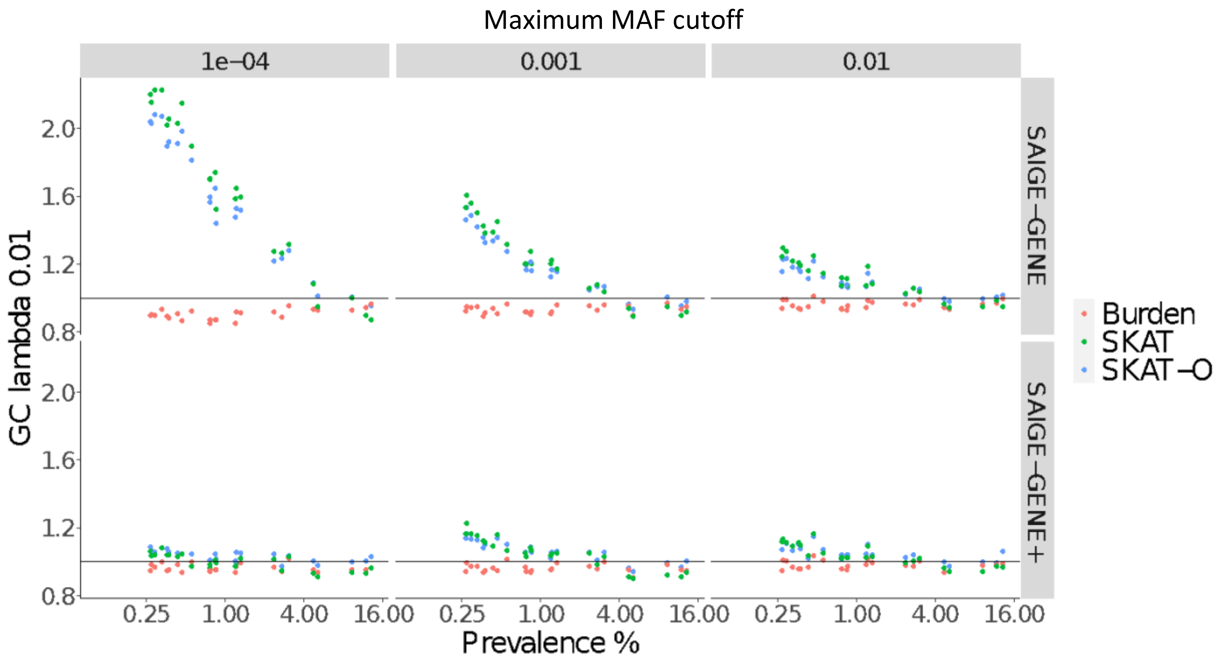

**Supplementary Figure 4. Quantile-quantile plots for Burden, SKAT[3], and SKAT-O[6] tests p-values for simulated phenotypes with prevalence 10%, 1%, and 0.3% based on the UKBB WES data under the null hypothesis.** A. Using SAIGE-GENE. B. SAIGE-GENE+, which collapses ultra-rare variants with MAC ≤ 10 prior to the gene-based association tests. The tests were performed for 18,372 genes with missense and loss-of-function variants with three different maximum MAF cutoffs: 1%, 0.1%, and 0.01%.

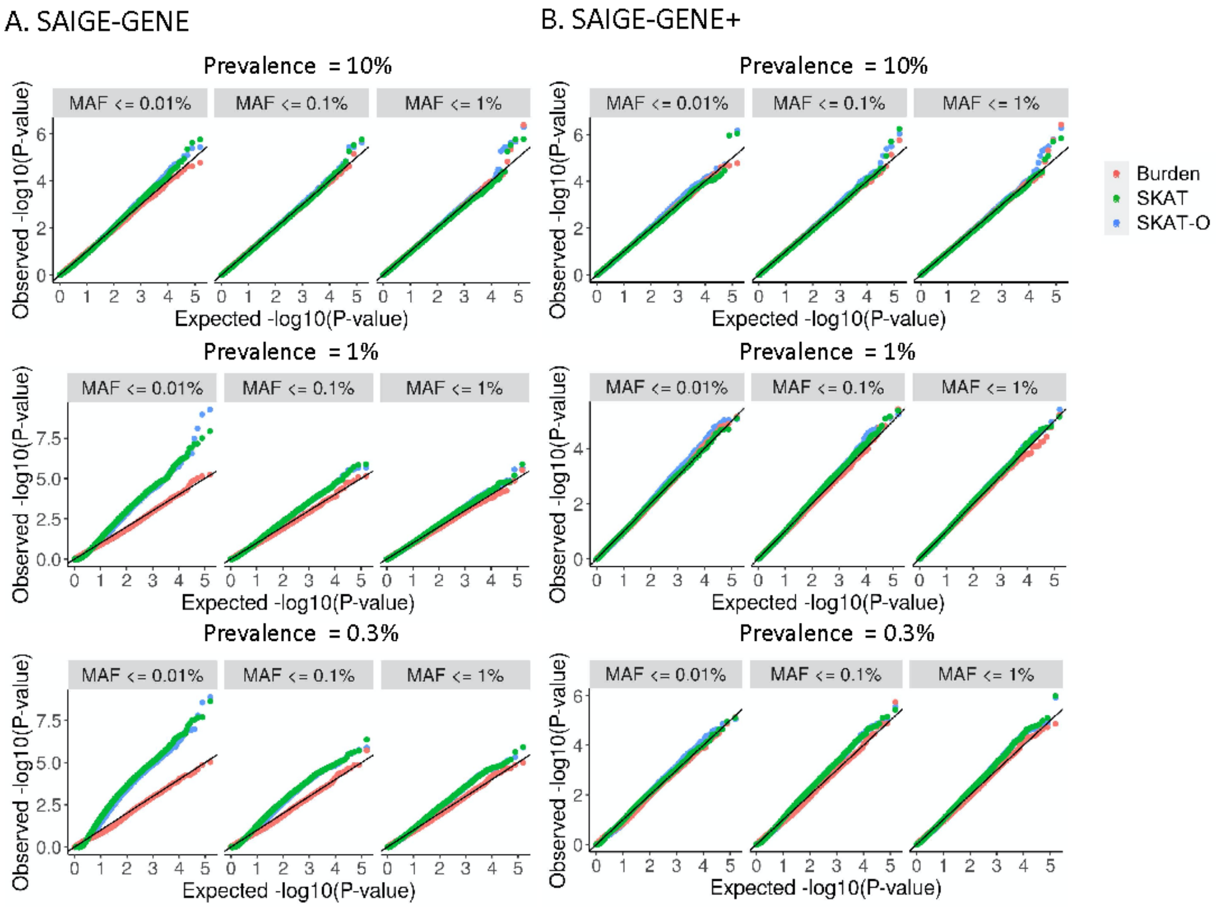

**Supplementary Figure 5.** Histogram of number of genetic variants (missense and LoF) tested in each gene with maximum MAF 1% before and after collapsing the ultra-rare variants with MAC <= 10.

A. All genes. B. genes with number of markers ≤ 500 before collapsing.

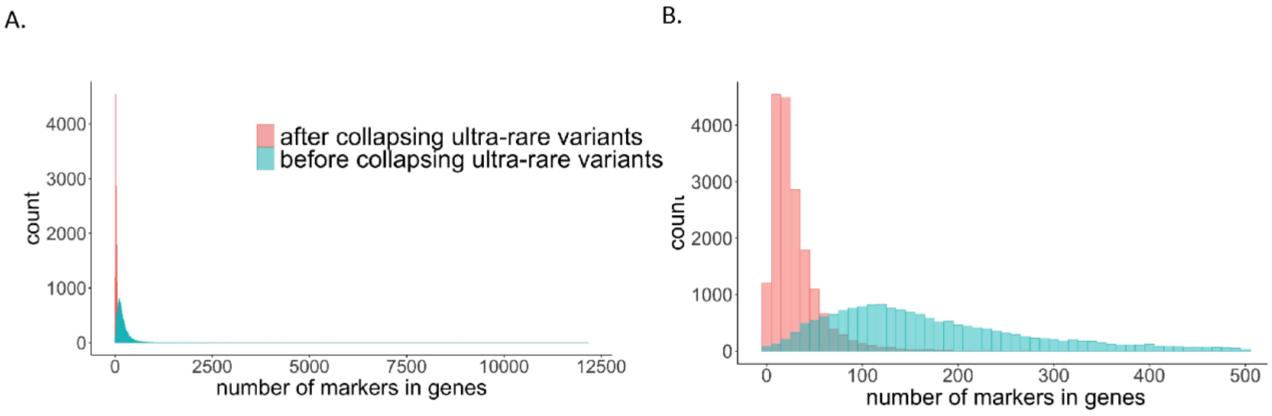

**Supplementary Figure 6.** Computation cost of the Step 2 in SAIGE-GENE+ with and without collapsing the ultra rare variants by sample sizes (*N*) for gene-based tests for 18,372 genes with three maximum MAF cutoffs: 1%, 0.1%, and 0.01% and three variant annotations: LoF only, LoF + missense, and LoF + missense +synonymous. In total around 165,348 tests were run for each data set.

Benchmarking was performed on randomly sub-sampled UK Biobank WES data with White British participants for glaucoma (1,741 cases and 162,408 controls). The reported run times and memory are medians of five runs with samples randomly selected from the full sample set using different sampling seeds. A. plots of the time usage as a function of sample size (N) B. plots of the maximum memory usage (for genes containing most variants) as a function of sample size (*N*). The x-axis is plotted on the log2 scale. C. scatter plots of the memory usage when N = 150,000 simulated with a random seed. We split the 165,348 tests into 133 chunks, each with ~150 genes. For each gene, 9 SKAT-O tests were conducted corresponding to three different MAF cutoffs and functional annotations followed by combining the p-values using the Cauchy combination Each dot in the plot is the maximum memory usage of a chunk among five runs with different random seeds.

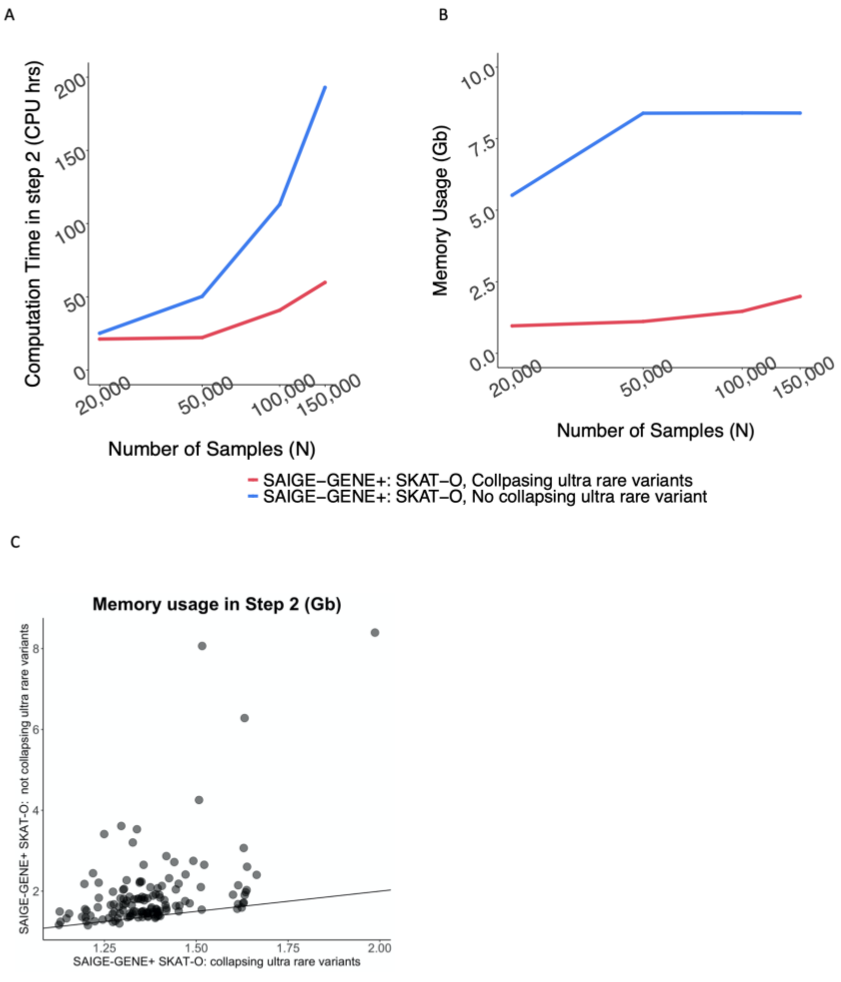

**Supplementary Figure 7.** Computation cost in SAIGE-GENE+ and REGENIE2 by sample sizes (*N*) for gene-based tests for 18,372 genes with three maximum MAF cutoffs: 1%, 0.1%, and 0.01% and three variant annotations: LoF only, LoF + missense, and LoF + missense +synonymous. In total 165,348 tests were run for each data set.

Benchmarking was performed on randomly sub-sampled UK Biobank WES data with White British participants for glaucoma (1,741 cases and 162,408 controls). The reported run times and memory are medians of five runs with samples randomly selected from the full sample set using different sampling seeds. A. plots of the time usage and median memory usage in Step 1 as a function of sample size (N) B. plots of the time usage and median memory usage in Step 2 as a function of sample size (N). Note that singletons only were also included as a mask in the Burden tests in both methods for a fair comparison. SAIGE-GENE+ further automatically output the p-values by the Cauchy combination

A.

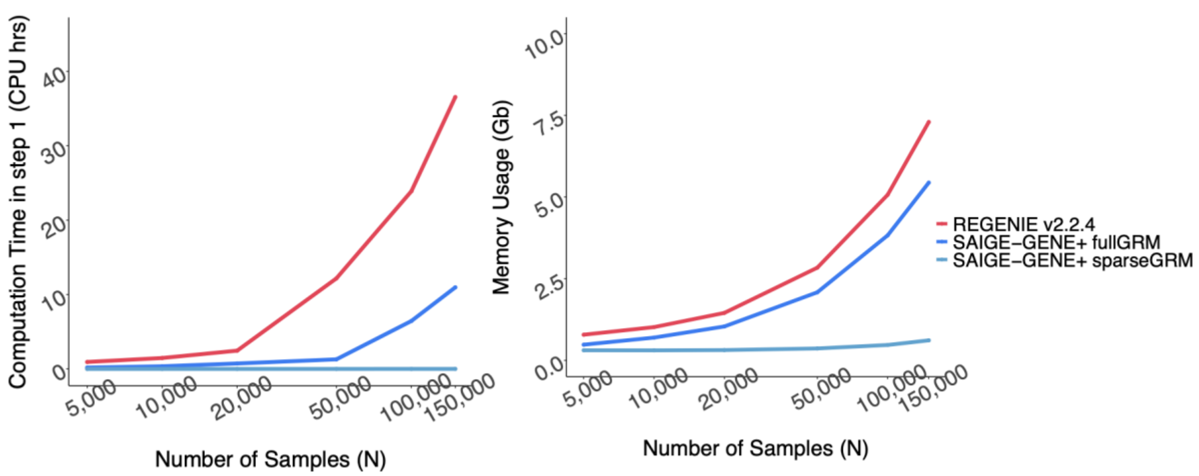


B.

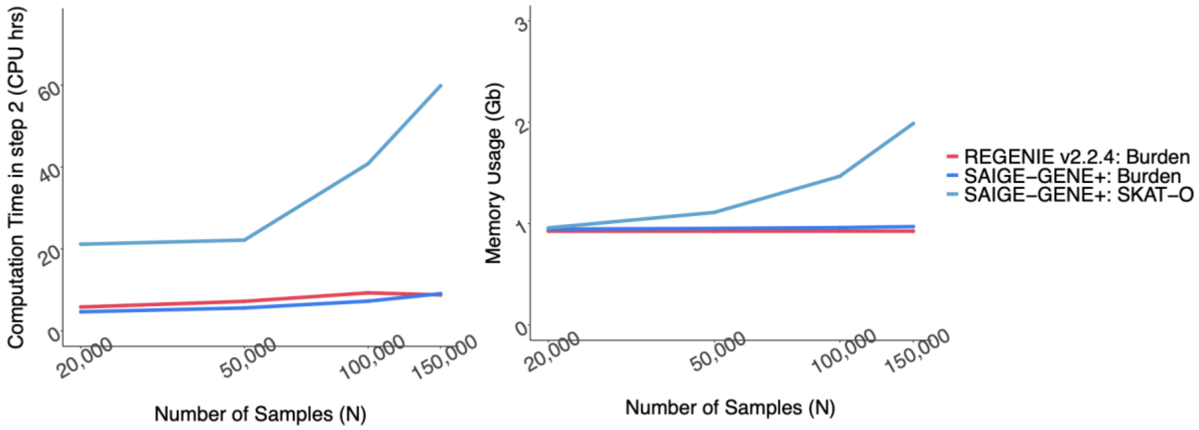

**Supplementary Figure 8. Scatter plots for association p-values by SAIGE-GENE+ and SAIGE-GENE in simulation studies for power evaluation.** Each plot is based on test results for 1,000 test sets (100 data sets, each of which includes 10 genes, see **Supplementary Table 5**). X-axis represents -log10 p-values without collapsing (SAIGE-GENE), and Y-axis represents -log10 p-values with collapsing (SAIGE-GENE+). The line in each plot represents the 45-degree line, so the dots above the line indicate more significant p-values from the collapsing. The details of different simulation settings are presented in **Supplementary Table 6**. A. All causal variants have the same effect direction. B. Causal variants have different effect directions (**Supplementary Table 6**).

A. Direction 1: All causal variants had negative effects

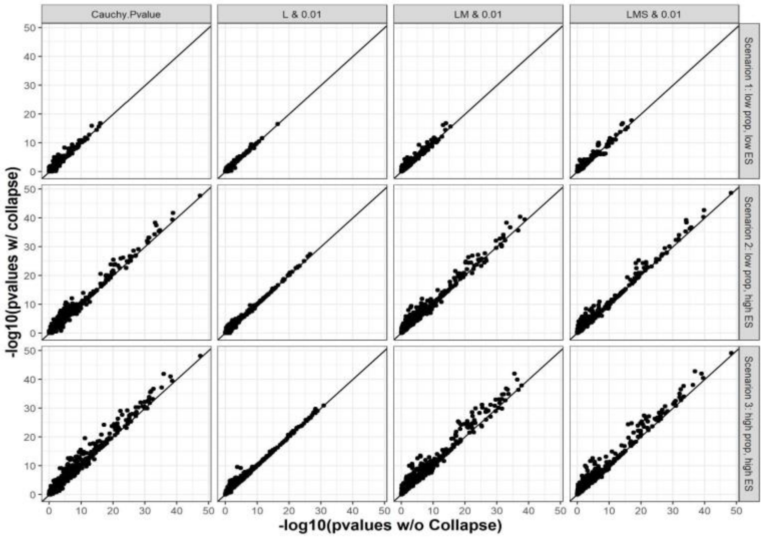


B. Direction 2: causal variants had different effect directions

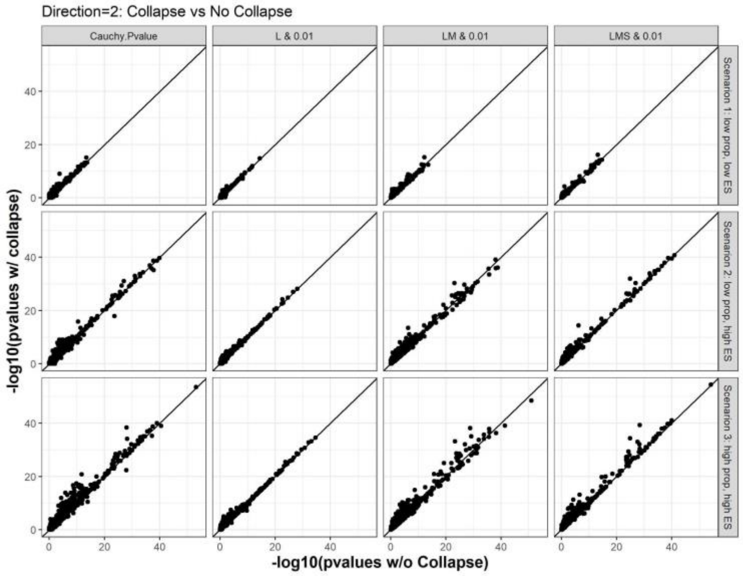

**Supplementary Figure 9. Scatter plots for association p-values of the Burden tests by SAIGE-GENE+ and REGENIE2.** The default weights (Beta(MAF, 1, 25)) for genetic variants are used in SAIGE-GENE+ and the "sum" mask was used in REGENIE2. A. All causal variants have the same effect. B. Causal variants have different effect directions (**Supplementary Table 6**).

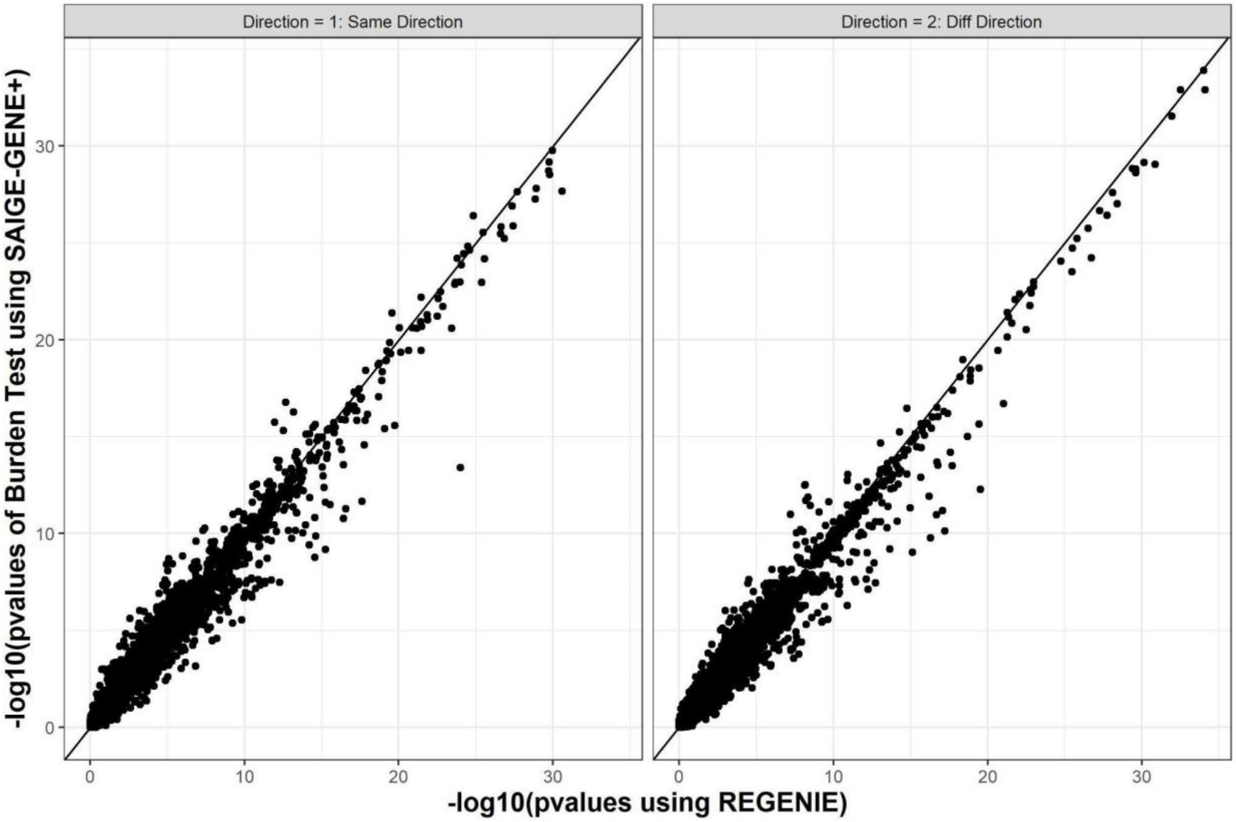

**Supplementary Figure 10. Scatter plots for association p-values in SAIGE-GENE+ using multiple MAF cutoffs and annotations and using a single cutoff and annotation.** In the former tests, three different function annotation combinations (L only, L+S, L+S+M) and three maximum MAF cutoffs (0.01%, 0.1%, 1%) were used. In the latter tests, one function annotation and maximum MAF cutoff (L+S+M with MAF <= 1%) are used. The details of different simulation settings are presented in **Supplementary Table 6**.

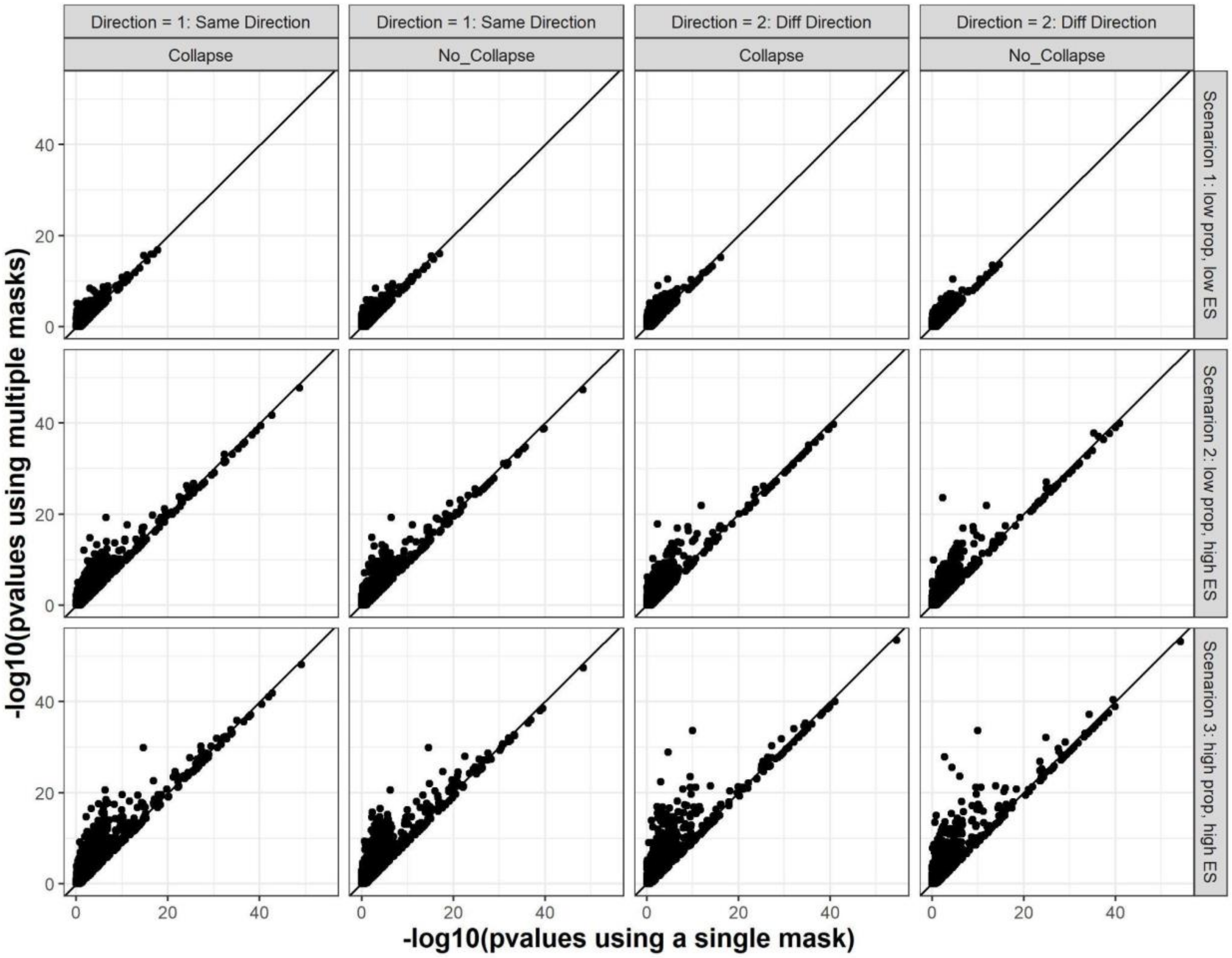

**Supplementary Figure 11.** Venn diagram of the number of associations identified (A) by different maximum MAF cutoffs (B) by different functional annotations

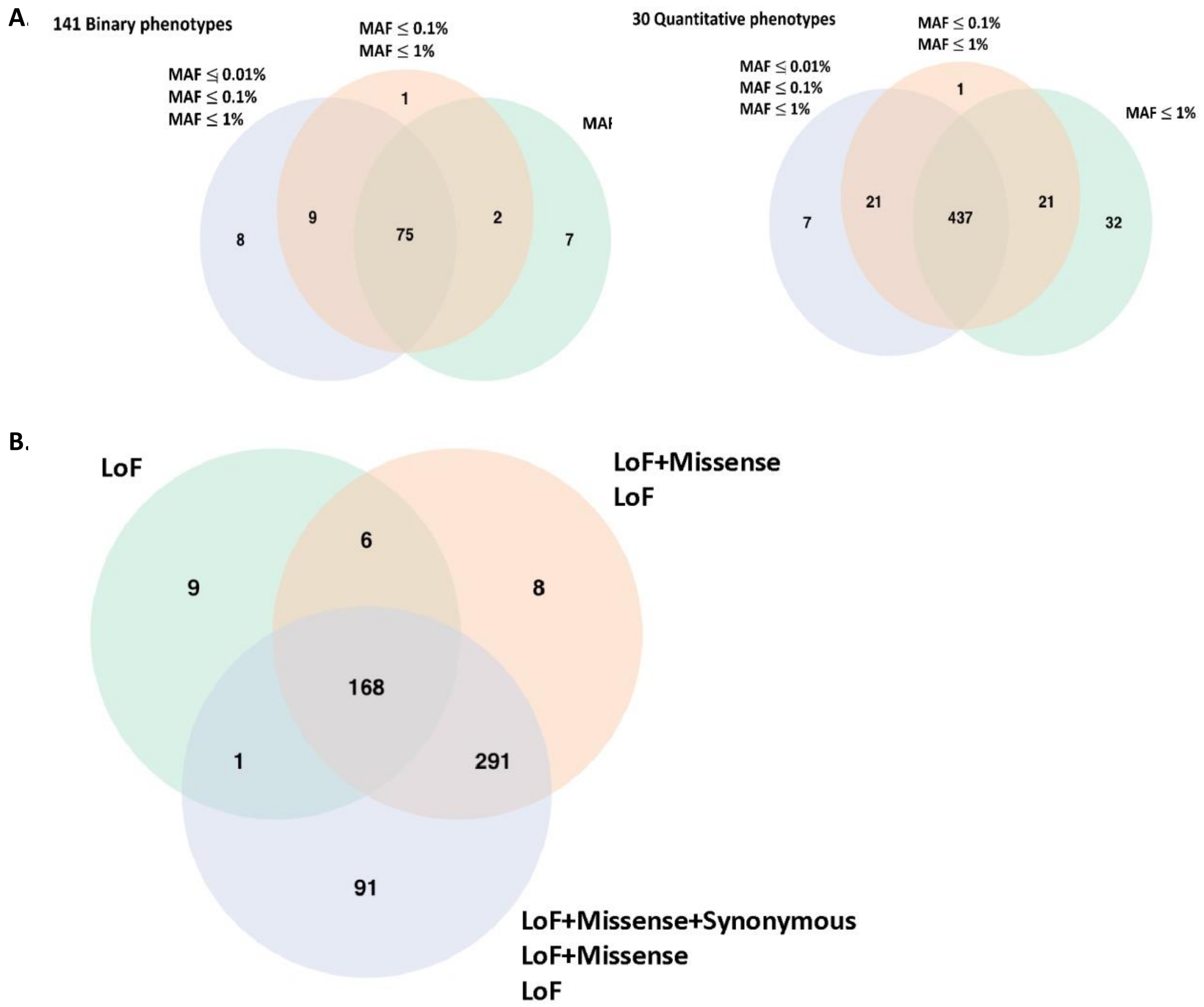

**Supplementary Figure 12.** Kinship coefficients (>= 0.05) in UKBB. A. all 408,910 samples B. 200,643 samples with whole exome sequencing data available

A.

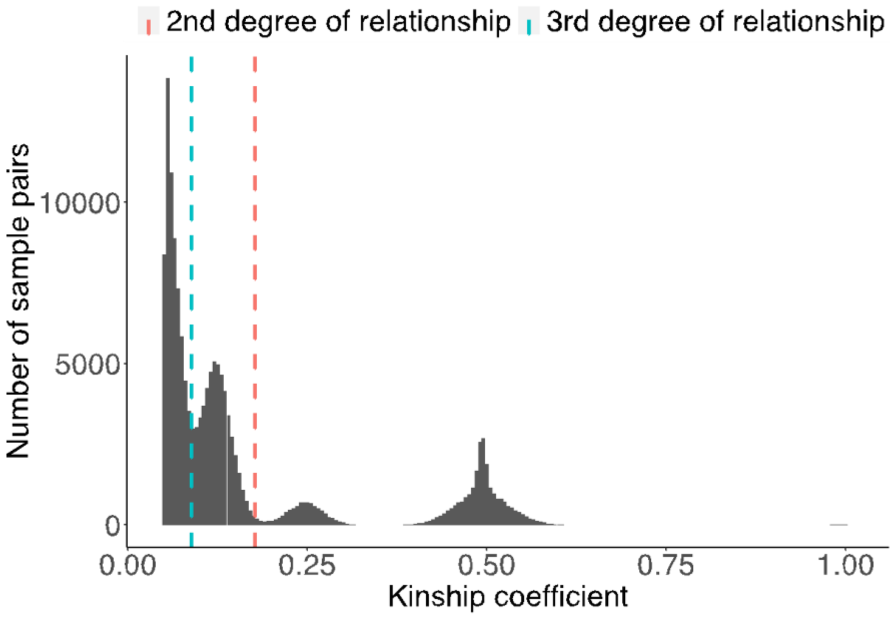


B.

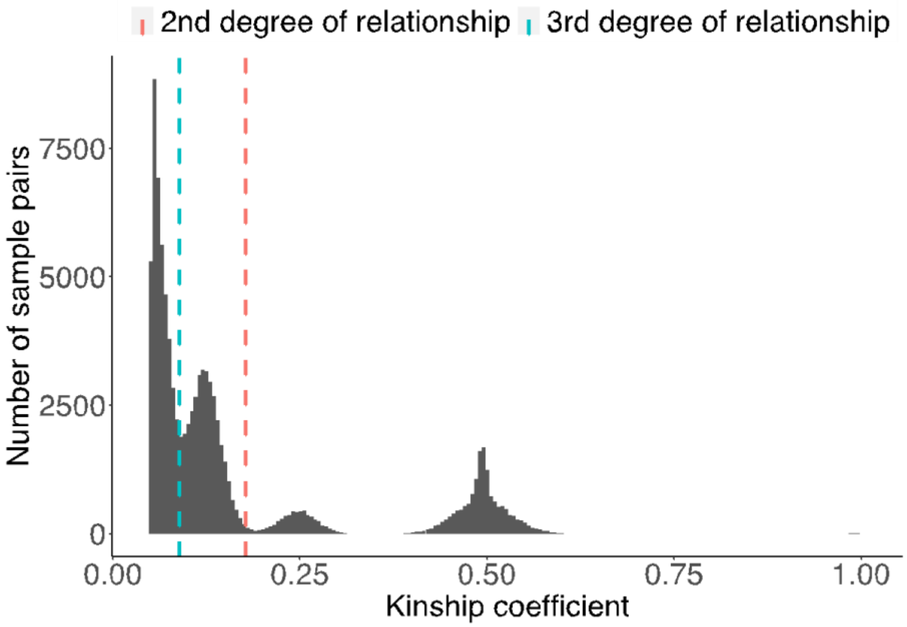

**Supplementary Figure 13**. Results with full GRM and sparse GRM for binary traits in UKBB WES data.

MAF <= 0.01%

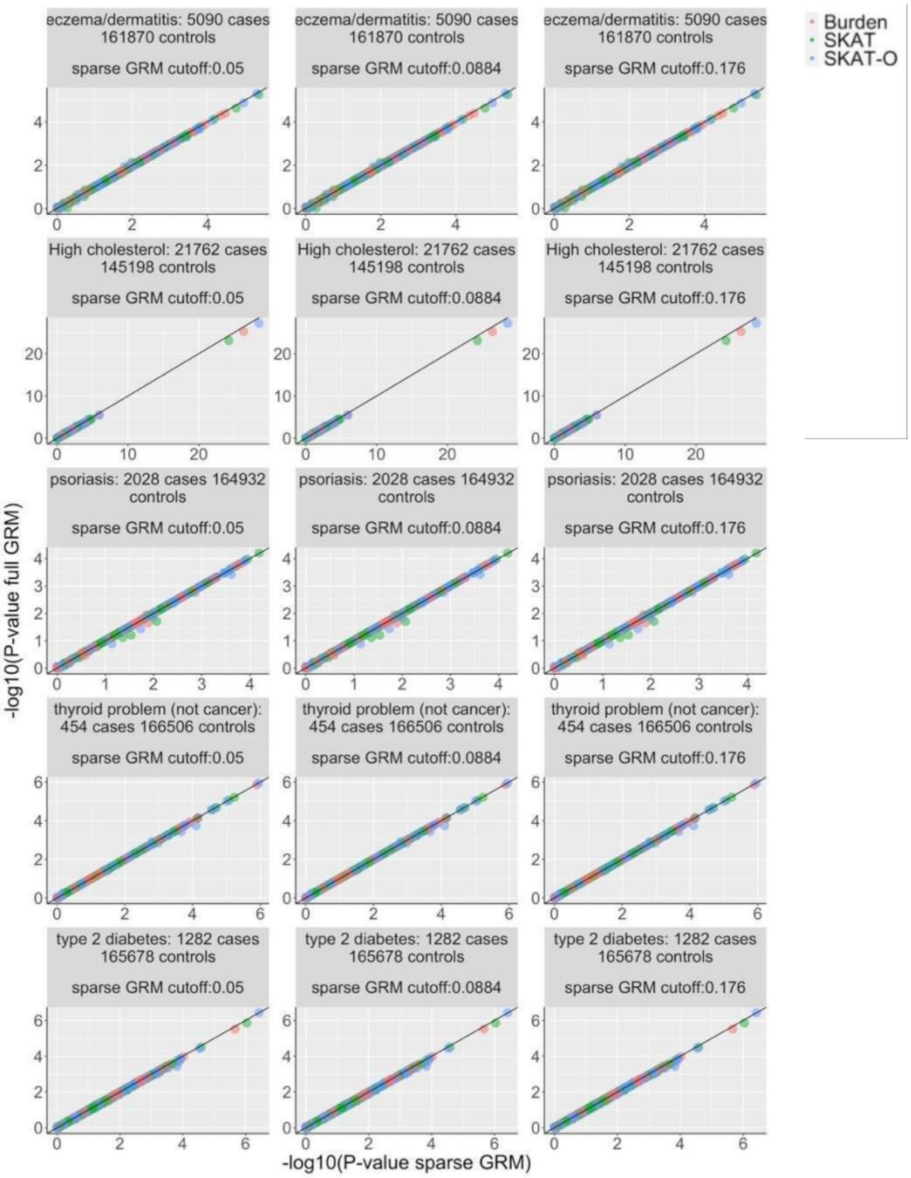

MAF <= 0.1%

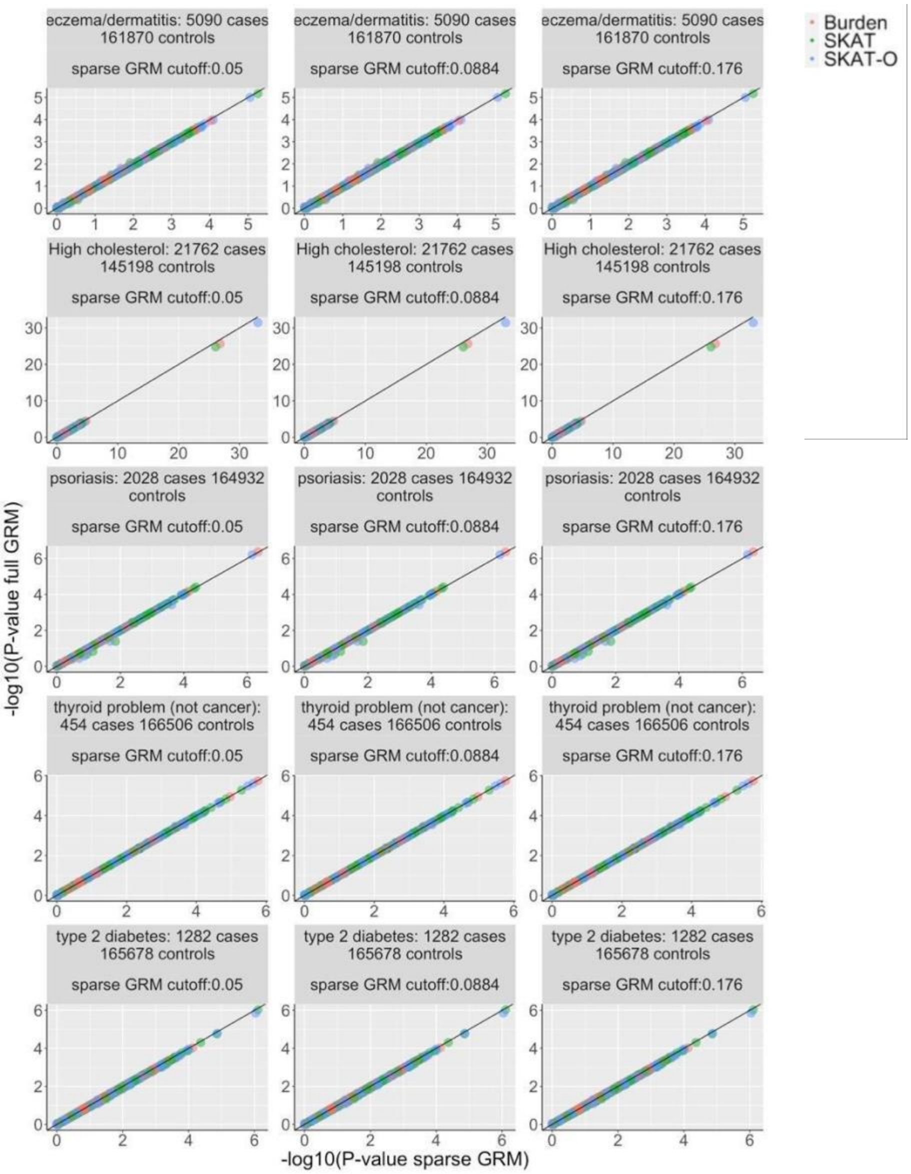
eczema/dermatitis: 5090 cases 161870 controls
sparse GRM cutoff:0.05
sparse GRM cutoff:0.0884
sparse GRM cutoff:0.176
High cholesterol: 21762 cases 145198 controls
psoriasis: 2028 cases 164932 controls
thyroid problem (not cancer): 454 cases 166506 controls
type 2 diabetes: 1282 cases 165678 controls
Burden
SKAT
SKAT-O
-log10(P-value full GRM)
-log10(P-value sparse GRM)

MAF <= 1%

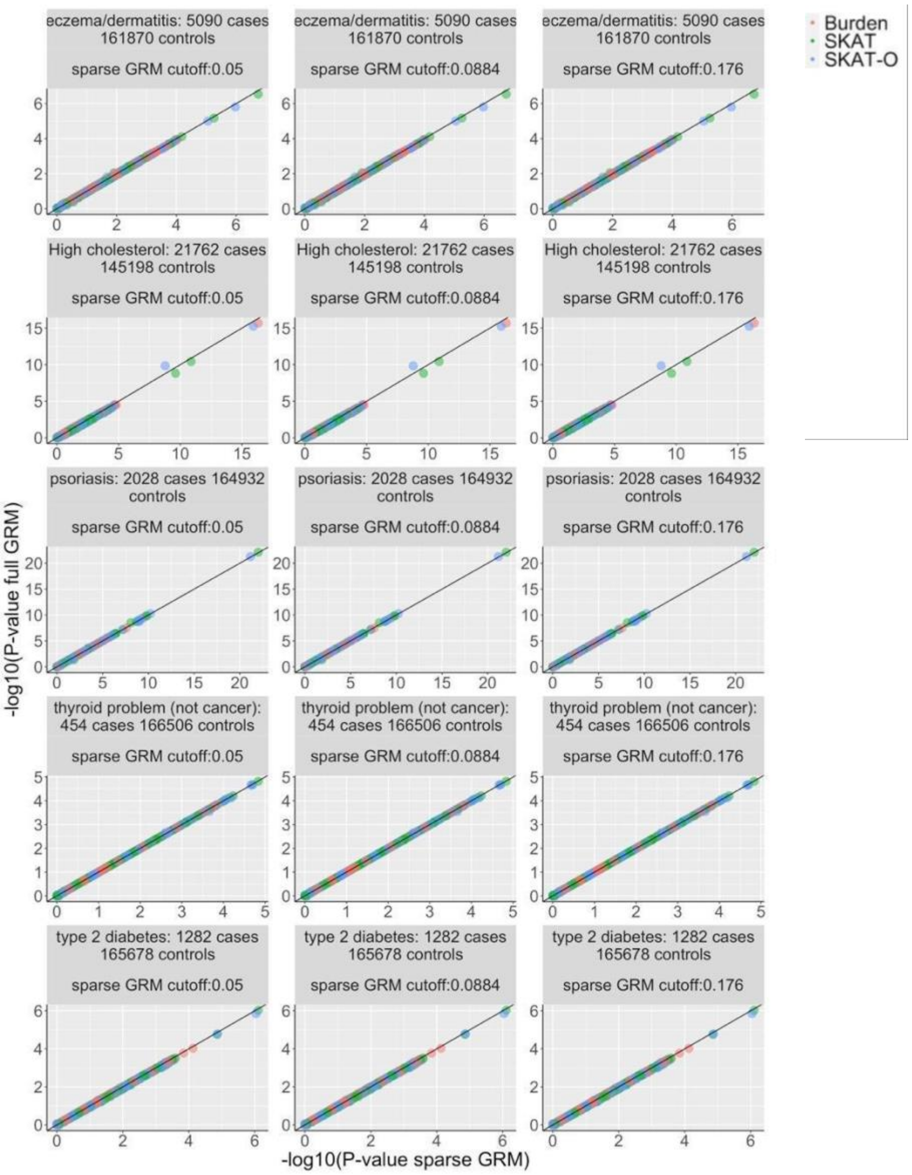
eczema/dermatitis: 5090 cases 161870 controls
sparse GRM cutoff:0.05
sparse GRM cutoff:0.0884
sparse GRM cutoff:0.176
High cholesterol: 21762 cases 145198 controls
psoriasis: 2028 cases 164932 controls
thyroid problem (not cancer): 454 cases 166506 controls
type 2 diabetes: 1282 cases 165678 controls
Burden
SKAT
SKAT-O
-log10(P-value full GRM)
-log10(P-value sparse GRM)

**Supplementary Figure 14.** Results with and full GRM and sparse GRM for quantitative traits in UKBB WES data

MAF <= 0.01%

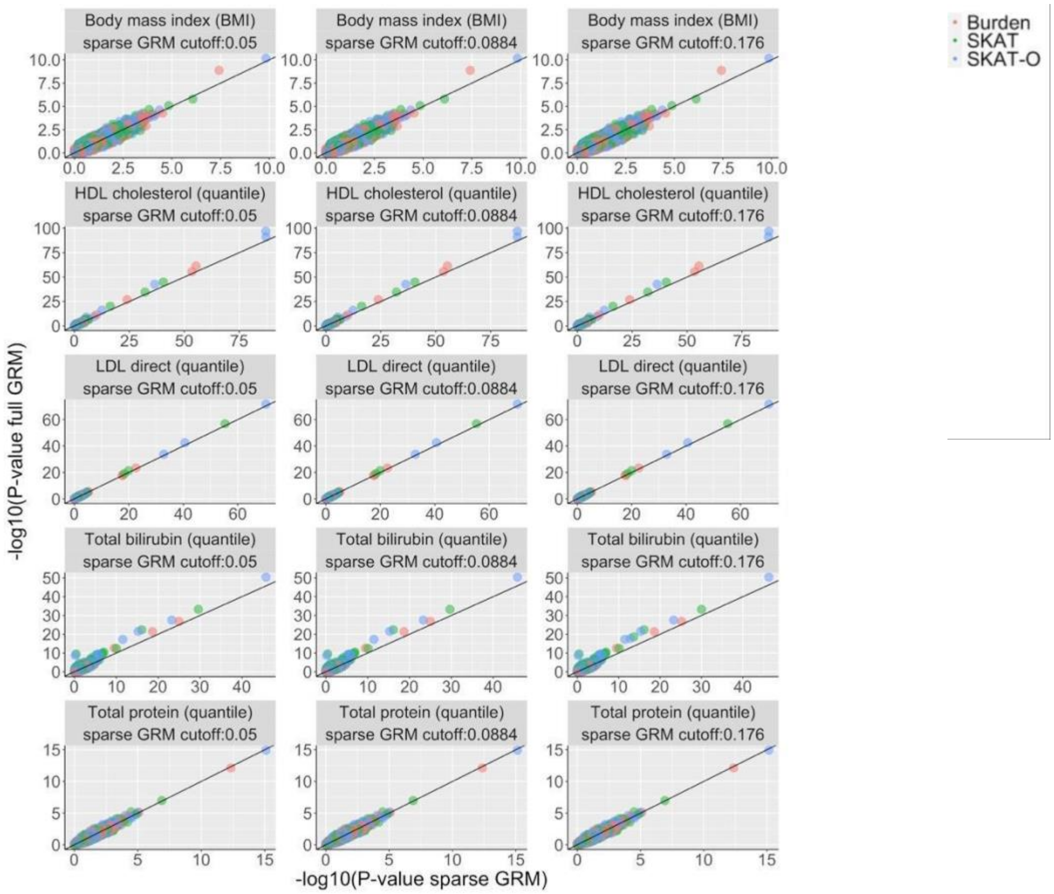

MAF <= 0.1%

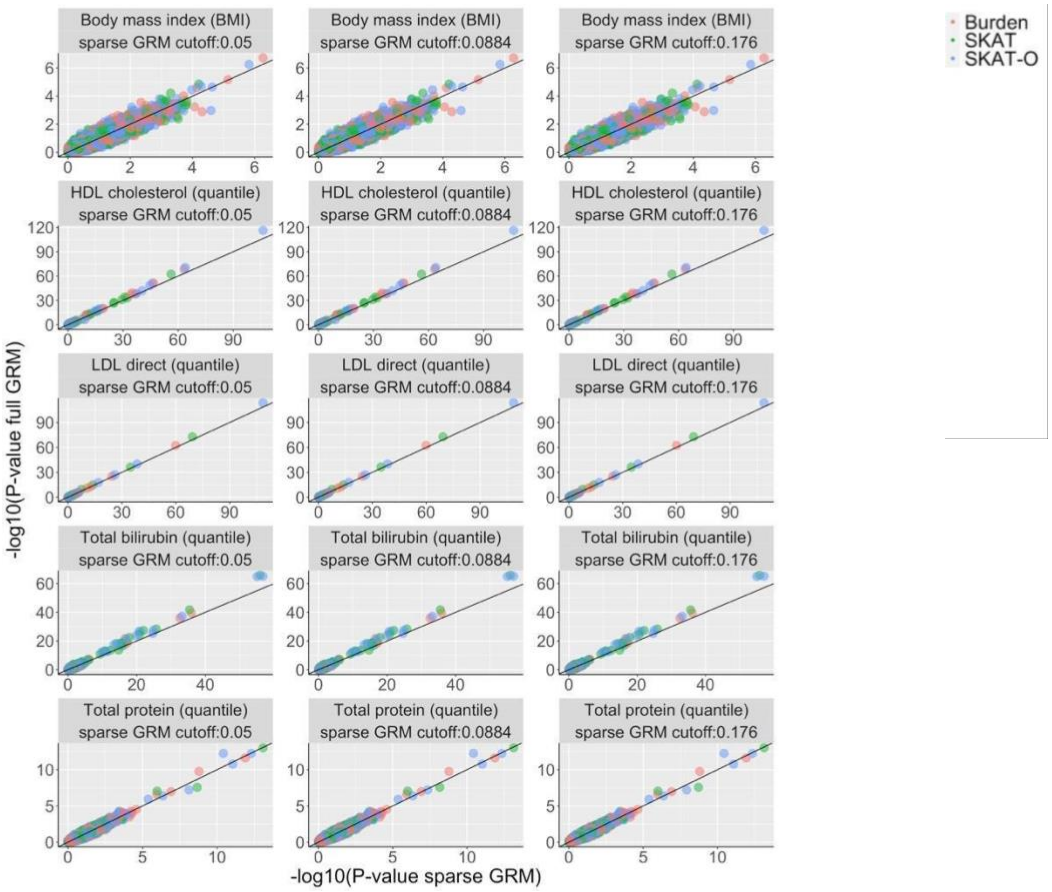

Body mass index (BMI)
sparse GRM cutoff:0.05
Body mass index (BMI)
sparse GRM cutoff:0.0884
Body mass index (BMI)
sparse GRM cutoff:0.176
HDL cholesterol (quantile)
sparse GRM cutoff:0.05
HDL cholesterol (quantile)
sparse GRM cutoff:0.0884
HDL cholesterol (quantile)
sparse GRM cutoff:0.176
LDL direct (quantile)
sparse GRM cutoff:0.05
LDL direct (quantile)
sparse GRM cutoff:0.0884
LDL direct (quantile)
sparse GRM cutoff:0.176
Total bilirubin (quantile)
sparse GRM cutoff:0.05
Total bilirubin (quantile)
sparse GRM cutoff:0.0884
Total bilirubin (quantile)
sparse GRM cutoff:0.176
Total protein (quantile)
sparse GRM cutoff:0.05
Total protein (quantile)
sparse GRM cutoff:0.0884
Total protein (quantile)
sparse GRM cutoff:0.176
-log10(P-value full GRM)
-log10(P-value sparse GRM)
Burden
SKAT
SKAT-O

MAF <= 1%

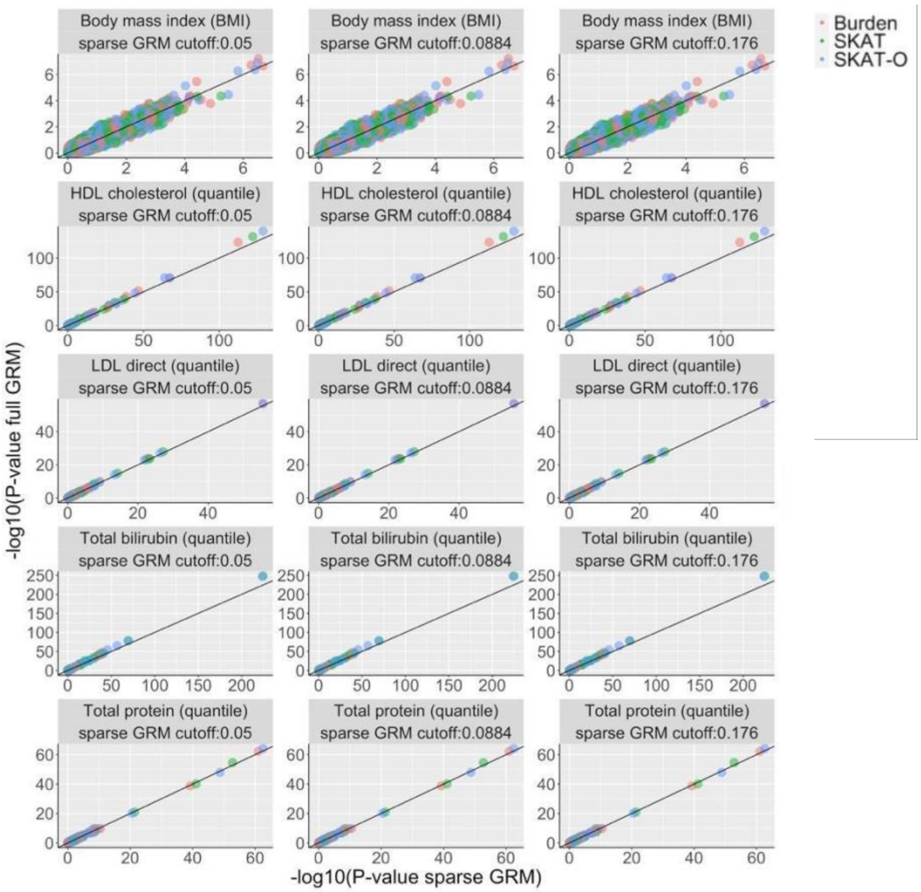

Body mass index (BMI)
sparse GRM cutoff:0.05
sparse GRM cutoff:0.0884
sparse GRM cutoff:0.176
HDL cholesterol (quantile)
LDL direct (quantile)
Total bilirubin (quantile)
Total protein (quantile)
Burden
SKAT
SKAT-O
-log10(P-value full GRM)
-log10(P-value sparse GRM)

**Supplementary Figure 15.** Collapsing ultra-rare variants with MAC $\leq$ 10.

A. No per-marker weights are provided in the group file by the user. The weights of the collapsed variant and other non-ultra-rare variants (MAC > 10) are calculated based on their MAFs from Beta distribution $w_j = Beta(MAF_j, a_1, a_2)$. By default, $a_1 = 1$, $a_2 = 25$.

| | Ultra-rare variants (MAC <= 10) | | | | | | | | | | | Collapsed (max of dosages) |
|---|---|---|---|---|---|---|---|---|---|---|---|---|
| Sample 1 | 0 | 0 | 0 | 0 | 0 | 0 | 0 | 0 | 0 | 0 | → | 0 |
| Sample 2 | 0 | 1 | 0 | 0 | 0 | 0 | 0 | 0 | 0 | 0 | → | 1 |
| Sample 3 | 1 | 0 | 0 | 0 | 0 | 0 | 0 | 0 | 1 | 0 | → | 1 |
| Sample 4 | 0 | 0 | 0 | 2 | 0 | 0 | 0 | 0 | 1 | 0 | → | 2 |
| Sample 5 | 0 | 0 | 0 | 0 | 1 | 0 | 1 | 0 | 0 | 0 | → | 1 |

*Weight of the collapsed variant*
$w = Beta(MAF, a_1, a_2)$

B. The per-marker weights are provided in the group file by the user

| | Ultra-rare variants (MAC <= 10) | | | | | | | | | | | Collapsed (max of the weighted dosages) |
|---|---|---|---|---|---|---|---|---|---|---|---|---|
| Sample 1 | 0 | 0 | 0 | 0 | 0 | 0 | 0 | 0 | 0 | 0 | → | 0 |
| Sample 2 | 0 | 1 | 0 | 0 | 0 | 0 | 0 | 0 | 0 | 0 | → | 0.2 |
| Sample 3 | 1 | 0 | 0 | 0 | 0 | 0 | 0 | 0 | 1 | 0 | → | max(0.1, 0.9) = 0.9 |
| Sample 4 | 0 | 0 | 0 | 2 | 0 | 0 | 0 | 0 | 1 | 0 | → | max(2*0.4, 0.9) = 0.9 |
| Sample 5 | 0 | 0 | 0 | 0 | 1 | 0 | 1 | 0 | 0 | 0 | → | max(0.5, 0.7) = 0.7 |
| User-specified weight | 0.1 | 0.2 | 0.3 | 0.4 | 0.5 | 0.6 | 0.7 | 0.8 | 0.9 | 1 | | |
